# Is Cancer significant Comorbid Condition in COVID 19 Infected Patients? -A Retrospective Analysis Experienced in a Tertiary Care Center in Eastern India

**DOI:** 10.1101/2022.05.14.22275079

**Authors:** Kalyan K Mukherjee, Aniruddha Dam, Deepa Chakrabarti, Debasish Jatua, Saubhik Sengupta, Rita Dutta, Suparna Majumdar, Shyam Sundar Mandal, Biswarup Basu, Pratiti Bhattacharjee, Dattatreya Mukherjee, Sankar Sengupta, Jayanta Chakrabarti

**Affiliations:** Chittaranjan National Cancer Institute, Kolkata, India; International School, Jinan University, P.R China

**Keywords:** Cancer, COVID 19, Comorbidity, RT PCR, Cohort study, low-income group

## Abstract

**Objectives:** Patients with a history of active malignancy were initially thought to be at a higher risk of having COVID-19, although available data are conflicting due to economic stress, malnutrition, fear of hospitalization or treatment discontinuation. A cohort-based study was undertaken in Indian regional cancer centre to understand cancer-covid link in patients.

**Study Design:** A total of 1565 asymptomatic patients were admitted based on thermal screening and evaluation from the screening form. The COVID 19 has been checked by RT-PCR method and the COVID 19 positive patients were transferred to government allocated COVID 19 hospital. The COVID 19 negative patients were transferred to general ward for further cancer treatment.

**Method:** Post COVID 19 testing, positive patients were transferred to COVID hospital and their outcomes were analyzed and correlated with patient’s age gender and cancer stages.

**Result:** Out of 1565 patients, 54 patients (3.4%) tested positive. Most of the patients are in 45-59 years age group. As female patients admitted were more in number than males, so predominance of disease is higher in female. 3 patients were symptomatic after admission and 2 were severe and were admitted to the ICU with ventilations. 8 patients died in Cancer and one patient died in COVID 19.

**Conclusions:** As only 3.4% patients tested positive and only one patient out of 54 had died, so cancer is found not to be a comorbid condition towards COVID 19 patients in the Indian population studied.

## Introduction

In late December 2019 first case of SARS-CoV-2 named by World Health Organization was reported in Wuhan, China. On January 31, 2020, World Health Organization (WHO) declared COVID-19 as a “Public Health Emergency of International Concern (PHEIC)” and on March 11, 2020, it was propounded as a “pandemic”. The rapid spread of SARS-CoV-2, combined with a near-complete global lockdown, has weakened the healthcare systems. Lack of adequate health care infrastructure and human resources, serious supply-chain disruptions, and widespread fear among patients and health care workers have resulted in patient care and safety being compromised.

The virus is transferred from person to person via respiratory droplets (coughing or sneezing) as well as through direct contact with an infected patient or fomites nearby (1,2). It is suspected that patients with comorbid illnesses are more susceptible viral infection complications (3). Patients with a history of active malignancy, particularly in lung, were initially thought to be at a higher risk of having COVID-19 and experiencing COVID-19-related complications, according to early studies (4,5,6).

In COVID-19 disease, it has been observed that the M1 macrophages are activated, which are associated with macrophage-activating syndrome (MAS), cytokine storm, lymphopenia, damage of the endothelium, and increase in intravascular blood coagulation. On the other hand, in cancer, M2 macrophages are activated that suppress the immune response and also simultaneously help in tumor progression. As a result of immunosuppression, the antiviral immune response is impaired that makes the cancer patients more susceptible to viral infections (7).

Previous research gathered and studied COVID-19 cases from 575 hospitals in 31 Chinese region. 18 of the total patients have a history of cancer, which may indicate a higher cancer rate than the Chinese population as a whole (0.9 percent vs. 0.29 percent). It also revealed that patients who had undergone chemotherapy or surgery were at a higher risk of having clinically severe events compared to patients who were not receiving these treatments for cancer. However, the interpretation of the findings in this study depended upon the small size of the cancer population (n=18), which acts as a limiting factor in drawing a conclusion (4).

A recent study of more than 1100 patients receiving treatment in oncology outpatient clinics in Germany reported that the vast majority of patients who tested positive for SARS-CoV-2 did not show any symptoms. Moreover, asymptomatic infections did not seem to impact the outcomes of further treatment such as chemotherapy. The findings — presented in a poster at the European Society of Medical Oncology (ESMO) Virtual Congress 2020 — conflicted with the belief that cancer patients in general tend to develop more severe forms of COVID-19.Demographic data obtained from the meta-analytical study based on 512 published papers and 13 studies, including a total of 3775 COVID-19 patients, 63 (1·66%) with Cancer and 3712 (98·3%) without cancer, was indicative of the fact that the cancer patients were elderly as compared to total data population (63 years versus 48·7 years). The mean age of 1862 patients in five studies were 51·8 years (SD.3). The gender distribution in 12 studies showed that 58% were males, while females constituted 42% of the population (8).

ASCO, CDC and others have launched guidelines to address various aspects of cancer care during the pandemic, including risk minimization; care prioritization of patients; health care team management; virtual care; management of patients with cancer undergoing surgical, radiation, and systemic therapy; clinical research; and recovery plans. Delay in surgery for potentially curable early tumors is a major conflict among the guidelines (9). Some of the guidelines followed in India based government-funded cancer hospitals were the establishment of “screening camps” outside the hospital to reduce patient visits, stringent restriction of relatives and friends in outpatient clinics and inpatient wards, the establishment of a fever clinic, and creation of isolation wards, rotation of staff to ensure a fallback option in case of mass quarantine (10).

The underlying immunosuppressive microenvironment due to dysfunctional immune cell production is a fundamental attribute of the malignancies that drives their pathogenesis (11). Comorbid conditions such as; Hypertension, Cardiovascular diseases, Chronic obstructive pulmonary diseases, Diabetes, Chronic kidney diseases, Cerebrovascular diseases are likely to decrease immunity, impair macrophage and lymphocyte function, and are associated with the pathogenesis of SARS-CoV-2 (12).

Previously reported observational studies showed hypertension (16-23%), cardiovascular disease (5-16.4%), diabetes (8-11.5%), cancer (3.9%), chronic kidney disease (2.4%), chronic obstructive pulmonary diseases (3.1%) and cerebrovascular diseases (3%) as the most frequently observed comorbidities among patients with SARS CoV-2. While hypertension (46.6%), diabetes mellitus (20.4%), cardiac disease (34.9%), cerebrovascular disease (9.1%), chronic liver disease (9.2 %), chronic kidney disease (10.8%) and chronic lung disease (14.7%) were reported as prevalent comorbidities among cancer patients (12). Patients with chronic respiratory disorders like chronic obstructive pulmonary disease, are prone to developing ARDS because of their lower resistance to the virus. In diabetic patients it induces inflammatory infection by causing the accumulation of activated innate immune cells in metabolic tissues which leads to the release of inflammatory mediators, especially IL-1β and TNFα (12).

From a systematic review of 31 studies and meta-analysis of 181,323 patients from 26 studies involving 23,736 cancer patients, it was observed that the most common type of cancer reported among COVID-19 patients were hematological malignancies (34.3%) followed by breast cancers (29.2%), lung cancers (23.7%), gastrointestinal malignancies (15.2%), prostate cancers (11.1%), gynecological cancers (9.6%), head and neck cancers (3.7%), brain tumors (3%) and other cancers (2.63%). Hematological malignancies had the highest pooled all-cause in-hospital mortality rate of (33.1%) followed by lung cancer at (28%), gastrointestinal malignancies at (19.8%), and breast cancer at (10.9%) (13).

It was further reported that the most common treatment modality reported in cancer patients affected with COVID-19 was chemotherapy (30.3%), followed by hormonal therapy (17.4%), targeted therapy (15.4%), radiotherapy (13.8%), immunotherapy (9.1%) and surgery (7.3%)(13).In response to the COVID-19 pandemic, the ASCO launched a Global Webinar emphasizing the need of protecting patients and health care teams from infections, delivery of timely and appropriate care, reduction of harm from the interruption of care, and preparation to handle a surge of new COVID-19 cases, complications, or comorbidities thereof (13). In this hospital based retrospective study 1565 patients have been enrolled from June 2020 to November 2020 out of which 54 (3.4%) patients were positive. During admission they were asymptomatic and it has been tried to make a correlation between COVID 19 and Cancer.

## Methods

### Study design

In a tertiary care hospital situated in eastern India, a total 1565 asymptomatic patients were enrolled in this study. The patients were admitted on the basis of thermal screening and evaluation from the screening form. High temperature, symptoms of COVID 19, travelling history in last 14 days are excluded from the study. All the patients (COVID 19 positive or Negative) were asymptomatic during admission and admitted in the 40 bedded isolation ward for treatment after RT PCR to check whether the patient has SARS-COV 2 positive or not. The COVID 19 has been checked by RT-PCR method and the COVID 19 positive patients have been transferred to government allocated COVID 19 hospital.

The COVID 19 negative patients are transferred to general ward from isolation ward for the further cancer treatment. Hospital followed standard procedures to limit any hospital infection among patients, health care professionals and caregivers through PPE, disinfectants etc. Institutional Ethical Committee of Chittaranjan National Cancer Institute has approved the study (IEC approval no: CNCI-IEC-KKM-11).

### RT-PCR based detection of COVID

BSL-2 labs were set up to handle patient samples and conduct RT-PCR testing. SARS nCoV-2 or Covid 19 testing was performed according to the guidelines of Indian Council of Medical Research (ICMR). Briefly, both oropharyngeal and nasopharyngeal swabs were collected and transported in Viral Transport Medium (VTM) to our laboratory. For each sample,140µl of VTM was used for extraction using HiPure Viral RNA Purification kit (HiMedia labs, India) according to the manufacturer’s instructions. The extracted RNA was measured by Nanodrop 2000 for estimating the quality of extracted RNA by 260nm/280nm. 5µl of extracted RNA was used for SARS nCoV-2 testing using different kits including Taqpath Covid19 multiplex real time RT-PCR kit (Thermo, USA), ViralDetect2 multiplex real time PCR kit for Covid 19 (Genes2Me, India), Diasure nCoV19 detection assay (GCC Biotech, India), MerilCov19 One Step RT-PCR Kit (Meril Diagnostics, India) approved and supplied by ICMR to our laboratory. In all the kits, E gene, RdRP or Orf1ab, N gene were used for detection and RNAseP was used as a housekeeping gene. RT-qPCR was run according to the manufacturer’s guidelines. Positive, Negative and Inconclusive samples were reported according to the kit manufacturer’s guidelines based on the PCR run. Validation was performed biannually according to the ICMR guidelines by Intra-laboratory testing and by External Quality Assurance Program (EQAS) as provided to us by ICMR. All the samples for validation were in 100% concordance with the intra-laboratory report and the EQAS report. In general, sensitivity and specificity of the kits were not mentioned in the kit literature.

### Patient based observational study

#### Inclusion Criteria

1. Asymptomatic Cancer Patients with no high fever and no recent history of travelling within 14 days of hospital admission screening.
2. Age more than 18 years (18-75 years)

#### Exclusion Criteria

1. If any family member is COVID 19 positive.
2. Very advanced or Terminal Cases.

Basic health criteria of patients from physical measurements were carried out for hospital admission. Basic comorbid conditions for both cancer or covid were considered.

### Statistical Analysis

Statistical Analysis was performed with help of Epi Info (TM) 7.2.2.2 EPI INFO is a trademark of the Centers for Disease Control and Prevention (CDC).Descriptive statistical analysis was performed to calculate the means with corresponding standard deviations (S.D.). Test of proportion was used to find the Standard Normal Deviate (Z) to compare the difference proportions. t-test was used to compare two means. p<0.05 was taken to be statistically significant.

## Result

### RT-PCR positivity

54 Patients (3.4 %) were found to be COVID-19 positive among 1565 patients admitted and associated with different degrees of viral load as observed by expression of marker genes.

### Correlation of Patient age, gender and cancer stages with treatment outcomes

Patient age, male-female ratio, comorbid features and cancer stages were studied and statistically correlated with outcomes in terms of COVID-19 or cancer (Table-1-6, Figure-I, II,).

**Table 1:** Age and Gender of Covid positive patients.

|  | Age |  |  |  | Gender |  |
| --- | --- | --- | --- | --- | --- | --- |
| | <30 | 30 - 44 | 45 - 59 | $\geq 60$ | Male | Female |
| **Covid positive** | 1 | 13 | 28 | 12 | 14 | 40 |
| **% Covid Positive patients** | 1.9% | 24.1% | 51.9% | 22.1% | 25.9% | 74.1% |

**Fig I:**
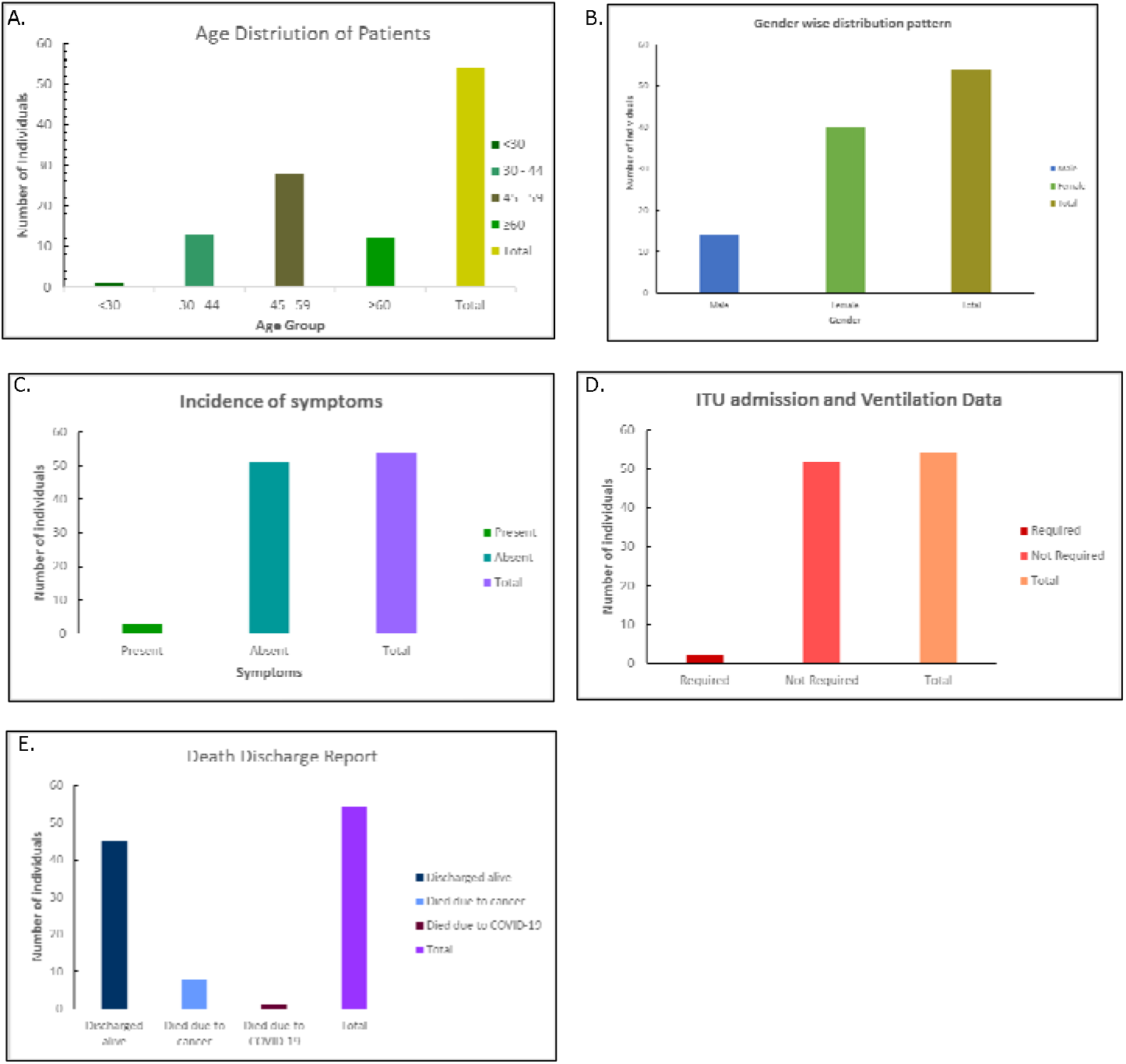
A: The Bar Graph shows the maximum affected age group is 45-59 years. In the hospital most of the patients got admitted with an age group of 40-60 years. B: Shows Females are more affected than male (Females are more admitted than Male). C: Shows that most of the cancer patients are asymptomatic during and after the admission. D: Shows majority of the patients didn’t go to moderate to severe stage, So, ventilation or ITU setup wasn’t a need. E: Shows most of the patient have discharged alive. Only 1 patient died due to COVID19 and 8 patients died due to Cancer.

**Fig II:**
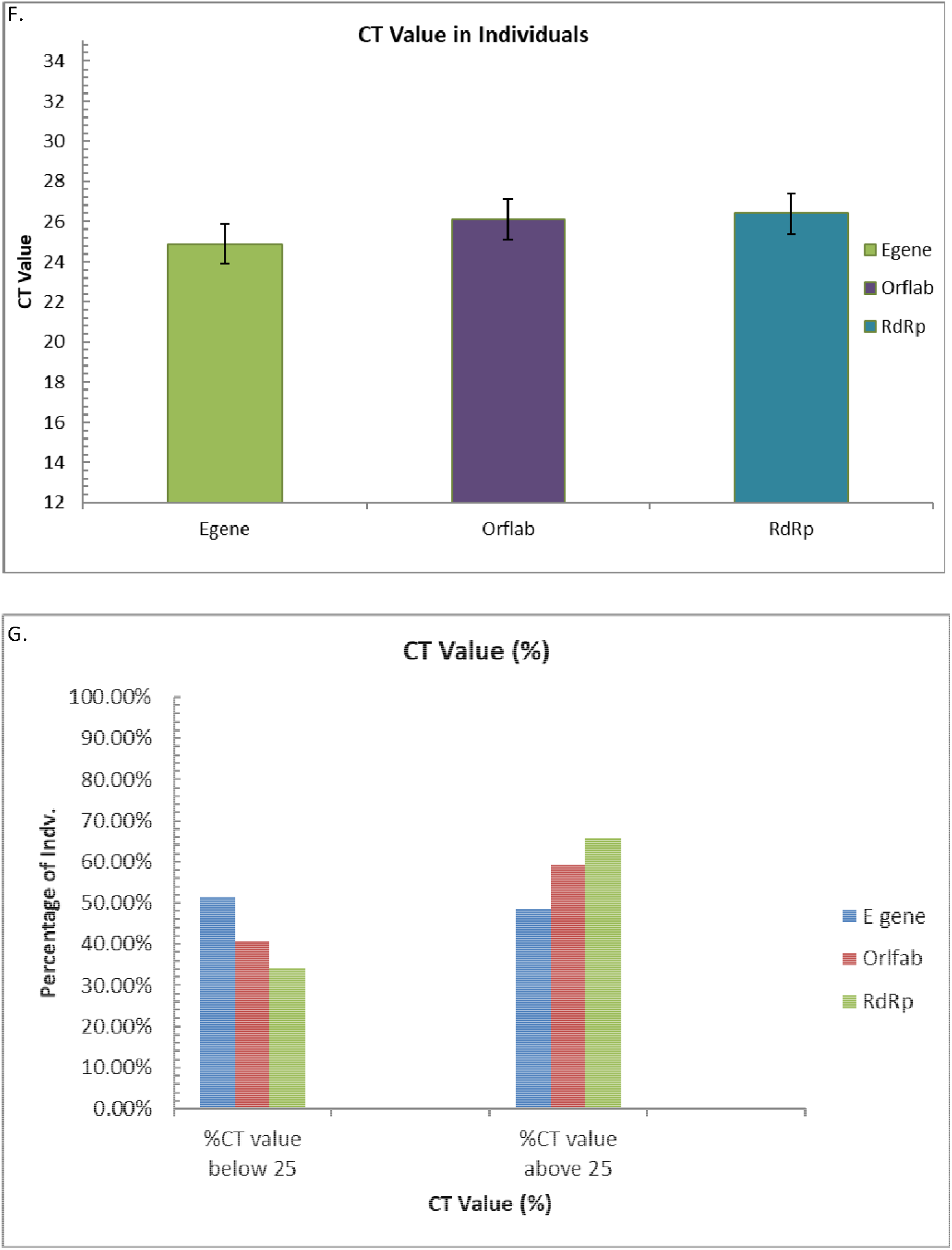
A: Comparative value of the three genes namely E gene, Orflab, RdRp used in CT value measurement among the patient study sample. B: Percentage of patients having CT value below or 25 and above 25

The mean age (± S.D.) of the patients was 50.38±10.82 with range 18 - 71 years and the median age was 49 years. Most of patients who got admitted in the hospital are at the age group of 40-60 years. (**Fig-IA)**74.0% of the patients were of age ≥45 years which was significantly higher than other ages (Z=6.78; p<0.0001). Thus, in this study COVID-19 infection was more prevalent among the patients with age≥45 years. (**Table-1)**

The ratio of male and female (Male: Female) was 1.0:3.6. Proportion of females (74.1%) was significantly higher than that of males (25.9%) (Z=6.78; p<0.0001). (**Fig-IB)** During admission more Female patient got admitted than male patient. (**Table-1)**

Though the mean age of the females was lower than that of the males, t-test showed that there was no significant difference between them (t_52_=1.06; p=0.30). However, females were at risk of having COVID-19 at younger age than males. (**Table-2)**

**Table 2:** Age and gender of the patients (t-test & p-value)

| Age<br>(In years) | Gender |  | t-test | p-value |
| --- | --- | --- | --- | --- |
|  | Male | Female |  |  |
| **Mean ± SD** | 52.57±10.6<br>2 | 49.62±10.92 | 1.06 | 0.30 NS |
| **Median** | 52.5 | 48.5 |  |  |
| **Range** | 32 - 69 | 18 - 71 |  |  |

**Table 3:**
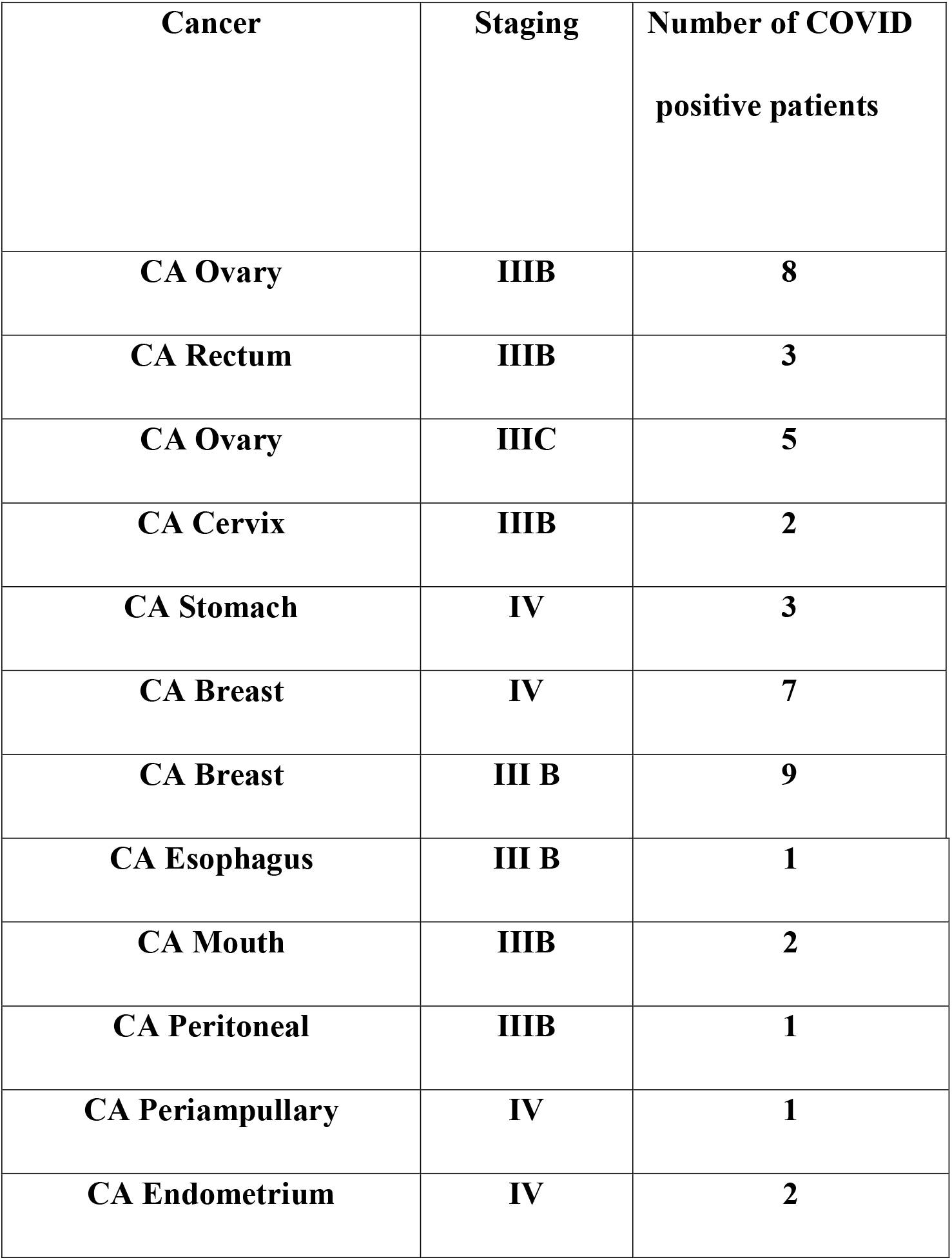
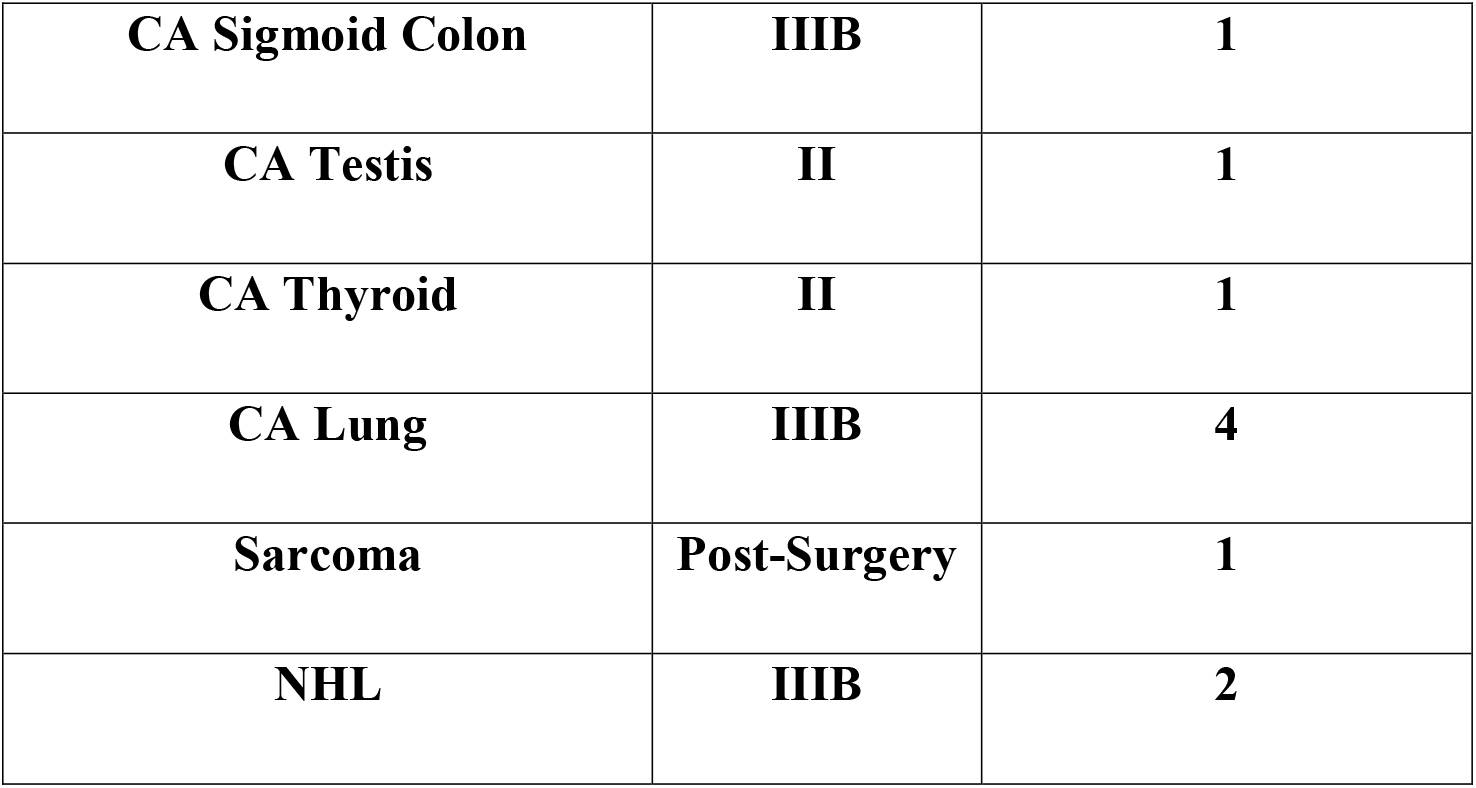
Types of Cancer and Staging and Number of Patients:

94.4% of the cases were asymptomatic which was significantly higher than symptomatic cases (5.6%) (Z=12.44; p<0.0001). (**Table-4), (Fig-IC**). In only 3.75 of the cases ITU admission was required. (**Table-4)**. In only 3.7% of the cases ventilation was required as respiratory support. (**Table-4), (Fig-ID)**.83.3%of the cases were discharged alive which was significantly higher than the patients died during treatments (16.7%) (Z=9.33; p<0.0001). A female patient aged 43 years died due to COVID-19. (**Table-5), (Fig-IE)**.

**Table 4:**
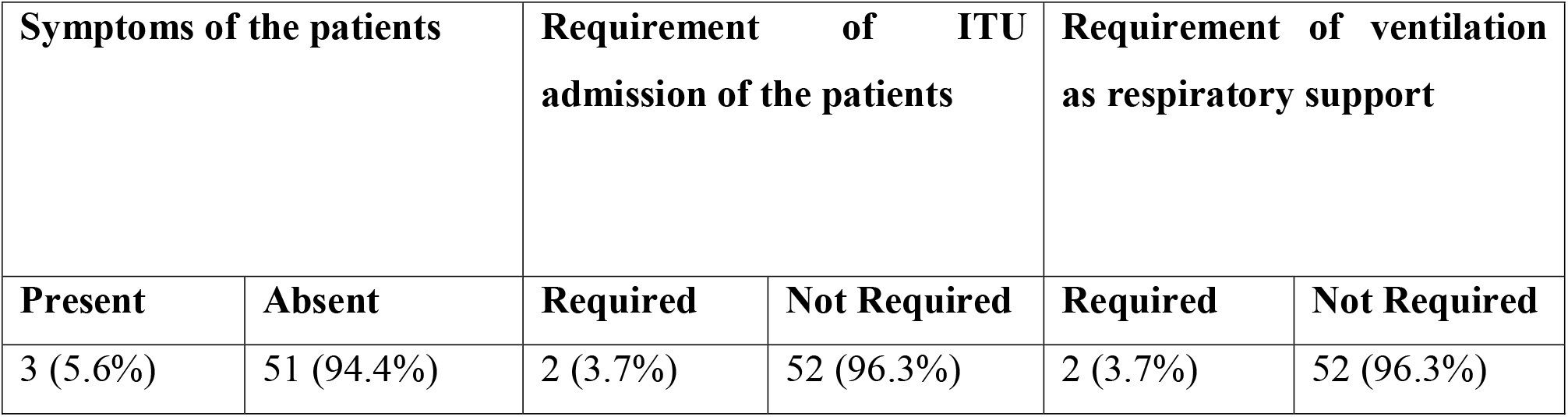
Symptoms, Requirement of ITU and Requirement of Ventilation of patients.

| **Symptoms of the patients** |  | **Requirement of ITU admission of the patients** |  | **Requirement of ventilation as respiratory support** |  |
| --- | --- | --- | --- | --- | --- |
| **Present** | **Absent** | **Required** | **Not Required** | **Required** | **Not Required** |
| 3 (5.6%) | 51 (94.4%) | 2 (3.7%) | 52 (96.3%) | 2 (3.7%) | 52 (96.3%) |

**Table 5:**
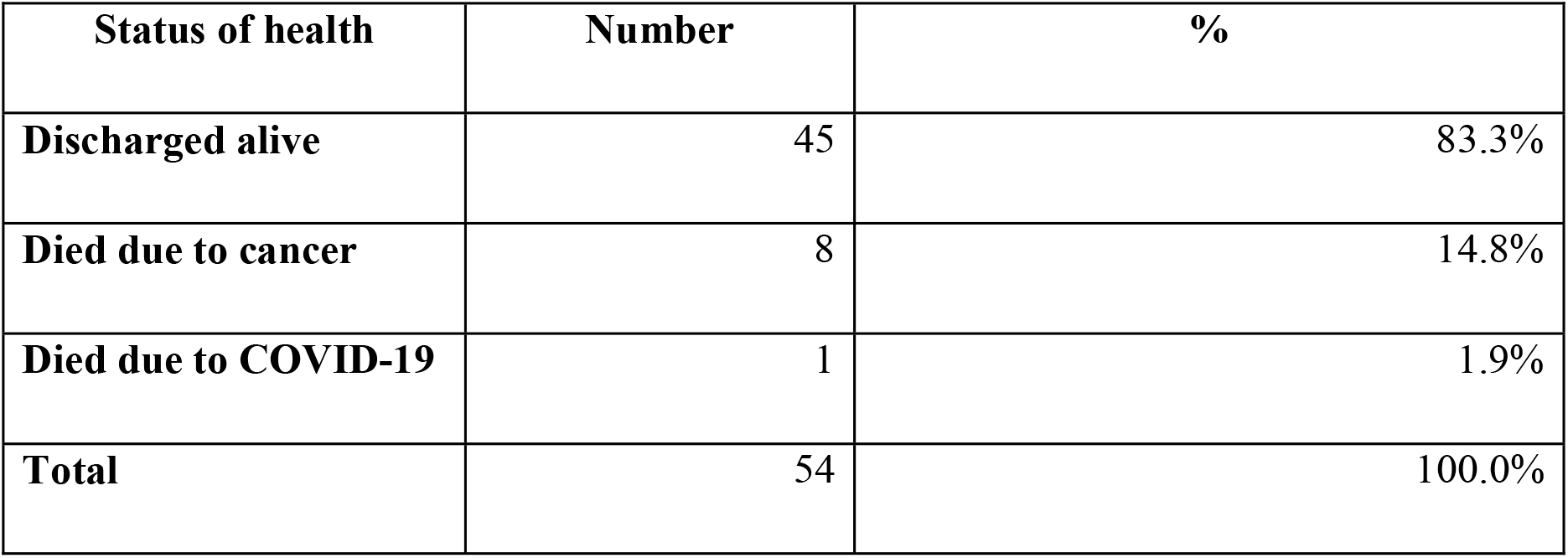
Status of health of the patients at discharge.

| Status of health | Number | % |
| --- | --- | --- |
| Discharged alive | 45 | 83.3% |
| Died due to cancer | 8 | 14.8% |
| Died due to COVID-19 | 1 | 1.9% |
| Total | 54 | 100.0% |

Comparative value of the three genes namely E gene, Orflab, RdRp used in CT value measurement among the patient study sample. Mean value of: E gene: 24.89189189, Orflab: 26.10810811, RdRp: 26.4. Standard deviation of: E gene: 5.440505, Orflab: 6.008253, RdRp: 5.882176 (**Fig-IIA)** Percentage of patients having CT value below or 25 and above 25 signifies the viral load. In the study it was see that percentage of patients having CT value above 25 was more. (**Fig-IIB) (Table-6)**.

**Table 6:**
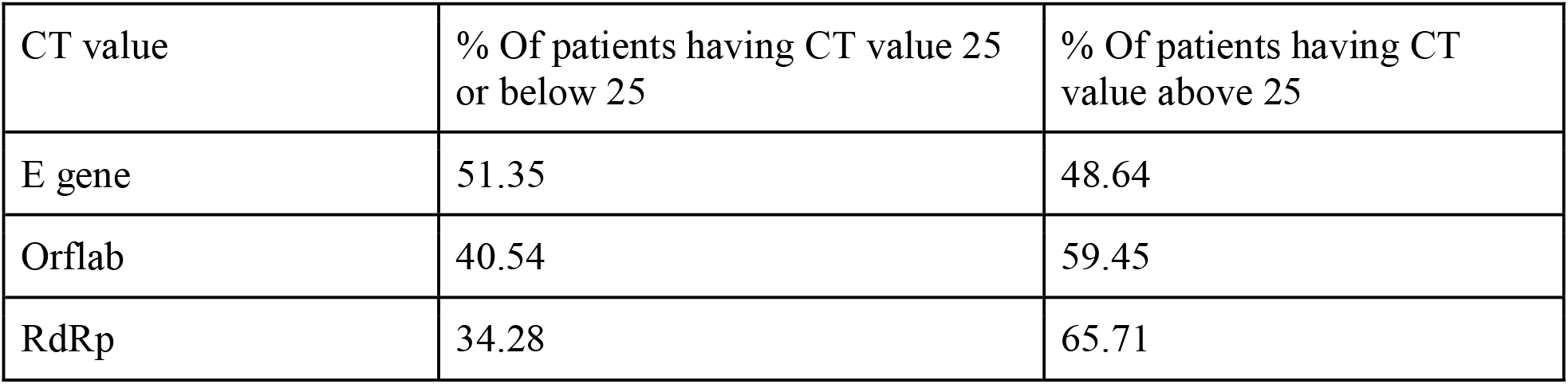
Percentage of patients having CT value 25 or below and above 25.

## Discussions

COVID-19 is strongly related with an inflammatory outburst, oxidative stress, and other pathophysiological abnormalities, which can exert enormous influence on the evaluation of cancer and treatment choices (14,15,16,17). In mild cases of COVID-19, there were substantial elevations in levels of numerous blood cancer indicators when compared to normal control participants, according to retrospective research. These cancer indicators were elevated even more in severe COVID-19 patients. (15). These changes might impact the positive and negative predictive values of a number of tumor-related biomarkers, making it more difficult to correctly assess and identify cancer diagnosis, disease progression, and treatment choices (18).

In our study, the 1565 patients were admitted in the hospital 40 bedded isolation ward through a screening process. In the isolation ward, COVID 19 testing was done in CNCI Rajarhat Campus and 54 patients (3.4%) were tested COVID 19 positive. All the patients were asymptomatic during admission.

COVID-19 testing prior to systemic treatment offers several advantages. To begin, the diagnosed patient can be spared potentially hazardous immunosuppressive therapy (chemotherapy, biologics, and immunotherapy). Secondly, for assessment, care, and isolation, the patient might be sent to an infectious disease expert. Third, contact tracing of the COVID-19-positive patient can be done and all the primary contacts can undergo COVID-19 testing to rule out asymptomatic COVID-19 infection. Contact tracing and isolation have both been found to be useful in breaking down chain transmission and fattening curves (19).

A Chinese retrospective analysis of 205 cancer patients with COVID-19 infection found that those who received treatment within four weeks after the beginning of symptoms had a high case fatality rate (20). The mortality was unrelated to cytotoxic chemotherapy or anticancer treatment in prospective observational research conducted in the United Kingdom on 800 individuals with cancer and symptomatic COVID-19 infection (21). Systematic strategizing, an environment that allows healthy disagreements, rapid multipronged implementation, willingness to change decisions on short notice, effective communication, and teamwork are all required to manage this pandemic, according to the Tata Memorial Hospital-COVID-19 working group (22).

This is one of the first study in India in a Government Setup where a large number of asymptomatic patients had undergone COVID 19 testing. This study has shown that only 3.6% of patients were COVID 19 positive. Testing adds to the workload of staff collecting specimens, increases the cost of personal protective equipment, creates a psychological burden for patients and their families if they test positive due to the delay in cancer treatment and the stigma associated with quarantine, and puts pressure on patients to be tested before each systemic therapy.

According to our study, this can be stated easily that Cancer is not a comorbid situation towards COVID 19 affected patients. Only 3.4% patients were tested positive and only 3 patients out of 54 were symptomatic, 2 patients were severely symptomatic and those patients had been admitted to ITU with ventilation support. Out of 54 patients, only one patient died in COVID 19. Another Indian Study have also shown similar scenario where 1.45% patients were COVID 19 positive. So, it can be stated that in Indian genetics and environment, COVID 19 is not a Comorbid situation.

## Conclusions

Our retrospective single center study in regional cancer center showed the following outcomes in cancer patients-

⍰ 54 patients out of 1565 have been COVID19 positive i.e. (3.4 %) COVID Positive
⍰ Over 45 years old patients are more venerable on the basis of this study result
⍰ Females are more affected than male.
⍰ Majority of patients (94.4%) where asymptomatic and 5.6% patients were symptomatic after the admission
⍰ ITU admission was not needed for 96.3% patients and ITU admission was done in 3.75% patients. These 3.75% patients were in ventilation.
⍰ 83.3% patients were discharged alive, 14.8% patients died due to cancer and 1.9% patients died due to COVID 19

Limitations of our study may be -most of the patients admitted in the age group of 40-60 years and female patients admitted were more in number than male patients. Our single center study has limitations of small cohort, so this study may be undertaken in larger cohort considering different demographic locations of East India and pooled data may be generated. Our study cohort represented that cancer is not a comorbid condition in COVID-19 infected patients where other comorbid conditions remaining almost same.

## Data Availability

All data produced in the present study are available upon reasonable request to the authors

## Declaration

We are sincerely thankful to everyone who have helped us in this study. We are also thankful to patients and their family members for their cooperation.

## Specific author’s contribution

Conceptualization: Dr Kalyan K Mukherjee;

Covid control Strategy: Dr. Aniruddha Dam, Dr. Deepa Chakrabarti, Dr. Debasish Jatua, Dr. Saubhik Sengupta,

RT-PCR Lab: Dr. Saubhik Sengupta, Dr. Sankar Sengupta

Data collection: Smt. Rita Dutta

Radiology Support: Dr. Suparna Majumdar

Statistical Study: Dr. Shyam Sundar Mandal, Dr. Dattatreya Mukherjee

Scientific writing: Dr. Biswarup Basu, Ms. Pratiti Bhattacharjee, Dr. Dattatreya Mukherjee

Institutional Support: Dr. Sankar Sengupta, Dr. Jayanta Chakrabarti

## Ethical Approval

Ethical Committee of Chittaranjan National Cancer Institute has approved the study. IEC Ref: CNCI-IEC-KKM-11

